# Optimizing Input Selection for Cardiac Model Training and Inference: An Efficient 3D CNN-based Approach to Automate Coronary Angiogram Video Selection

**DOI:** 10.1101/2024.04.22.24306184

**Authors:** Shih-Sheng Chang, Behrouz Rostami, Gerardo LoRusso, Chia-Ha Liu, Mohamad Alkhouli

## Abstract

**Background:** Research leveraging deep learning (DL) for medical image analysis is increasingly using dynamic coronary angiography from cardiac catheterizations to train neural networks. Yet, an efficient, automatic method to select appropriate dynamic images for training is still largely missing.

**Methods:** We developed DL models using 254 coronary angiographic studies from the Mayo Clinic. We utilized two state-of-the-art Convolutional Neural Networks (CNN: ResNet and X3D), to identify low quality angiograms through binary classification (satisfactory/unsatisfactory). Ground truth for the quality of the input angiogram was determined by two experienced cardiologists. We validated the developed model in an independent dataset of 3,208 procedures from 3 Mayo sites.

**Results:** 3D-CNN models outperformed their 2D counterparts, with the X3D model achieving superior performance across all metrics (AUC 0.98, precision 0.86, and sensitivity 0.89). The 2D models processed the video clips faster than 3D models. Despite having a 3D architecture, the X3D model had lower computational demand (2.56 GMAC) and parameter count (2.98 M) than 2D models. When validating models on the independent dataset, slight decreases in AUC and sensitivity were observed but accuracy and specificity remained robust (0.88 and 0.89, respectively for the X3D model).

**Conclusion:** We developed a rapid and effective method for automating the selection of coronary angiogram video clips using 3D-CNNs, potentially improving model accuracy and efficiency in clinical applications. The X3D-S model demonstrates a balanced trade-off between computational efficiency and complexity, making it suitable for real-life clinical applications.

---

**A**s machine learning (ML) integrates into cardiology, there’s a growing trend of using invasive X-ray video angiograms from cardiac catheterization for ML-driven analyses. This approach allows for evaluating left ventricular systolic function^1^, coronary vessel stenosis^2^, and myocardial ischemia^3,4^. Video images represent a valuable source of data for ML models but require careful selection and preparation before effectively utilized. The performance of ML models in this context heavily relies on selecting the appropriate DICOM images. Nevertheless, the process is complex due to the variability in image acquisition, the non-negligible background noise, and the high frequency of irrelevant or non-diagnostic images (e.g., short cine to verify puncture site or catheter location). These issues hinder the performance of the best ML models. Additionally, the image quality can be affected by other factors like poor catheter engagement, implanted devices (e.g., pacemakers), or suboptimal filling of the contrast, reducing image clarity and model accuracy ^5,6^. Hence, there is an unmet need to identify efficient techniques to select the optimal images for model training and testing.

Automated methods have already emerged to improve selection efficiency, such as automated keyframe image selection and object detection technology^2,7^. However, these methods were developed from still images and might not be suitable for angiographic clip selection. Other methods use object detection to label items in the cine images such as heart, arteries, catheters, and non-cardiac structures (e.g., ribs, diaphragm, pacemaker, clip, or tube). However, these method demand large databases and substantial labor to set up and annotate the images and may not perform well for rarely encountered objects.

Due to the lack of effective tools for selecting high-quality images from video coronary angiograms for ML purposes, we developed a new workflow that utilizes advanced Convolutional neural networks including 2D (ResNet 2D) and 3D (ResNet 3D and X3D) architectures. ResNet 2D models focus on spatial features in images, while ResNet 3D models are designed to capture temporal information^8,9^. The X3D model, a 3D CNN architecture, enhances video processing efficiency and accurately predicts left ventricular function in a prior study^1^. By integrating this selection tool into the data preprocessing step, we aim to enhance the accuracy of downstream models and improve the overall inference process.

## Methods

### Data Selection

The utilized dataset of cardiac catheterization imaging was meticulously curated by enlisting adult patients who underwent coronary angiography at Mayo Clinic locations in Minnesota, Florida, and Arizona between January 1, 2010, and December 31, 2021. An institutional review board approval was obtained. Patients who did not consent to use their medical records for research were excluded. A preliminary selection of 254 cardiac catheterization examinations was conducted using a random sampling methodology to develop a high quality benchmark model. A set of 3,208 cardiac catheterization examinations was then used to validate the model. We also validated the performance of the best models in a larger dataset comprising 20,388 angiographic video clips.

### Annotation Criteria

We annotated individual video clips within each angiographic study rather than make binary selections at the study level to ensure that no potentially useful video clips for model training were accidentally omitted. Two board-certified cardiologists independently reviewed the angiogram video clips. The following criteria were utilized to classify video clips as unsatisfactory: (1) Non-coronary angiograms, such as angiograms of the aorta, left ventricle, or other structures; (2) Angiography images in which the coronary arteries are obscured due to improper catheter engagement, inadequate contrast filling, or partial recordings during percutaneous coronary interventions; (3) Angiography images of graft vessels in patients who have undergone coronary artery bypass surgery; (4) Angiograms in which the presence of foreign objects, including pacemakers or spinal implants, obstructs the coronary arteries, making them difficult for the model to discern. We carefully monitored each image to exclude only angiograms where the foreign objects significantly obstructed the visualization of most coronary arteries. This approach ensures quality assurance, model generalizability, without unnecessary video discarding.

### Model Training

We uniquely and randomly divided angiogram images based on Patient ID into training, validation, and test sets with proportions of 70%, 15%, and 15%, respectively. In this study, both 2D and 3D CNN architectures (ResNet-18, ResNet-152, and X3D-S) were employed to evaluate the quality of the angiograms^10–12^. These models were chosen due to their proven capabilities in processing visual information and extracting image features, which are particularly beneficial for video analysis.

DICOM images were converted to PNG and resized to ensure model compatibility—160×160 pixels for 3D CNNs and 299×299 pixels for 2D CNNs. We selected videos with a frame rate of 15 frames per second based on DICOM header information, ensuring consistent temporal resolution for analysis. To capture the most representative frames from angiogram videos, we selected 32 frames starting from the 16^th^ frame of each video clip. This range was chosen to avoid initial frames which often lack relevant content and final frames which can be affected with contrast washout. For the 2D CNN-based experiments, one frame per eight frames within this range was selected, providing comprehensive coverage of the video content. In contrast, with 3D-CNNs, we sampled every other frame, considering the minimal variation between consecutive frames and focusing on image quality assessment rather than detailed temporal relationships.

These models were trained using Python 3.10.12 and the PyTorch framework version 1.12.1. The 2D and 3D ResNet models underwent a training process spanning >100 epochs. We utilized the Adam optimizer, complemented by a step decay learning rate schedule (StepLR) with a step size of 30. The 2D ResNet model was configured with an learning rate of 5e-5, a weight decay factor of 1e-3, and a batch size of 32. Similarly, the 3D ResNet model was set with a learning rate of 1e-4 and similar weight decay and batch size values. The X3D-S model followed a similar strategy but differed in weight decay at 1e-4 and a batch size of 64. Besides optimizing hyperparameters, we used data augmentation methods like flipping, rotating, perspective changes, Gaussian blurring, and image normalization to mimic real clinical variations and aid the models to better adapt to actual clinical situations.

### Model Evaluation

The efficacy of our models in classifying angiogram images into satisfactory or unsatisfactory was assessed using the following metrics^13^ ^14^. The Area Under the Receiver Operating Characteristic curve (AUC) reflects the model’s discriminative ability by plotting sensitivity versus 1 – specificity, with higher values indicating superior performance. Sensitivity, the proportion of true positive cases correctly identified, is given by 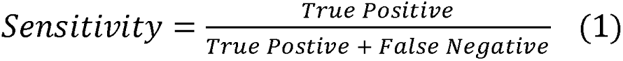. Specificity, representing the true negative rate, is defined as 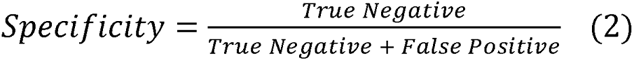. Accuracy representing the proportion of true results (both true positives and true negatives) in the total number of cases examined, is defined as 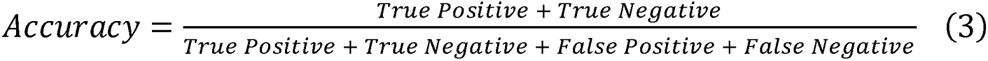. Precision, the ratio of true positive predictions to all positive predictions, is 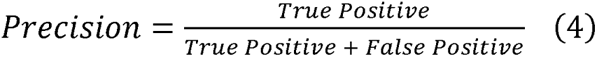. Lastly, the F1 Score, a harmonic mean of precision and recall, is 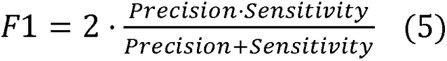. Accuracy encapsulates the overall effectiveness of the models, precision is vital for reducing false positive diagnoses, and the F1 score balances the trade-off between precision and recall, which is particularly relevant in the face of class imbalance.

To assess the model’s complexity and efficiency, we used the ptflops package 0.7.2.2 to calculate the multiply–accumulate operation (MAC)^15^. Parameters refer to the total count of weights, bias terms, and other parameters processed during the model’s training, with the unit of measurement being millions (M). A smaller number of parameters indicates a lower computational cost. MACs serve as a widely-used measure of computational complexity especially in models that rely heavily on linear algebra operations like CNNs^10,16,17^. In addition, we used the frames per second (fps) metric, an indicator used in previous studies^16^, to represents the number of frames transmitted per second, which reflects the real-time nature of the ML model. Each model’s time required to process a single video clip was reported for comparative analysis at the video level. These additional metrics are crucial for understanding the feasibility of deploying the ML models in clinical environments where computational efficiency can be limited. We performed model training on the mForge platform, utilizing NVIDIA V100 GPUs supported by the collaborative infrastructure between Mayo Clinic and the National Center for Supercomputing Applications, enhanced by a 100Gb/s InfiniBand network.

## Result

### Dataset Composition and Quality Classification

The study’s characteristics are presented in **Table 1**. It consists of 857 video clips deemed satisfactory and 180 video clips deemed unsatisfactory. The mean age of the participants were 62.7 and 64.7 years in the satisfactory vs. unsatisfactory groups. The percentage of male patients was 57.3% in the satisfactory group and 60% in the unsatisfactory group.

**Table 1.** Dataset Characteristics for Model Development.

|  | Satisfactory | Unsatisfactory |
| --- | --- | --- |
| Number of video clips | 857 | 180 |
| Number of exams | 150 | 104 |
| Number of patients | 135 | 96 |
| Age (year, mean ± SD) | 62.7±13.2 | 64.7±12.1 |
| Male gender (%) | 57.3 | 60.0 |

### Performance Metrics Across CNN Architectures

In the model development **Table 2** summarizes the relative effectiveness of various CNN architectures. Among the 2D models, the ResNet-18 achieved an AUC of 0.89, while the ResNet-152 achieved an AUC of 0.93 (**Figure. 1**). Using the 3D architectures, the ResNet-18 and ResNet-152 models achieved significantly higher AUCs of 0.95, and 0.97 (**Figure. 1**). The X3D-S architecture achieved an AUC of 0.98 along with high scores in accuracy of 0.96, precision of 0.86, and specificity of 0.98 (**Figure. 1**). The F1 scores across the models, indicative of a balance between precision and recall, ranged from 0.63 to 0.87, with the 3D models generally outperforming the 2D counterparts. **Figure 2** illustrates the adeptness of the X3D-CNN model in distinguishing between satisfactory and unsatisfactory angiograms within the test set.

**Figure 1.**
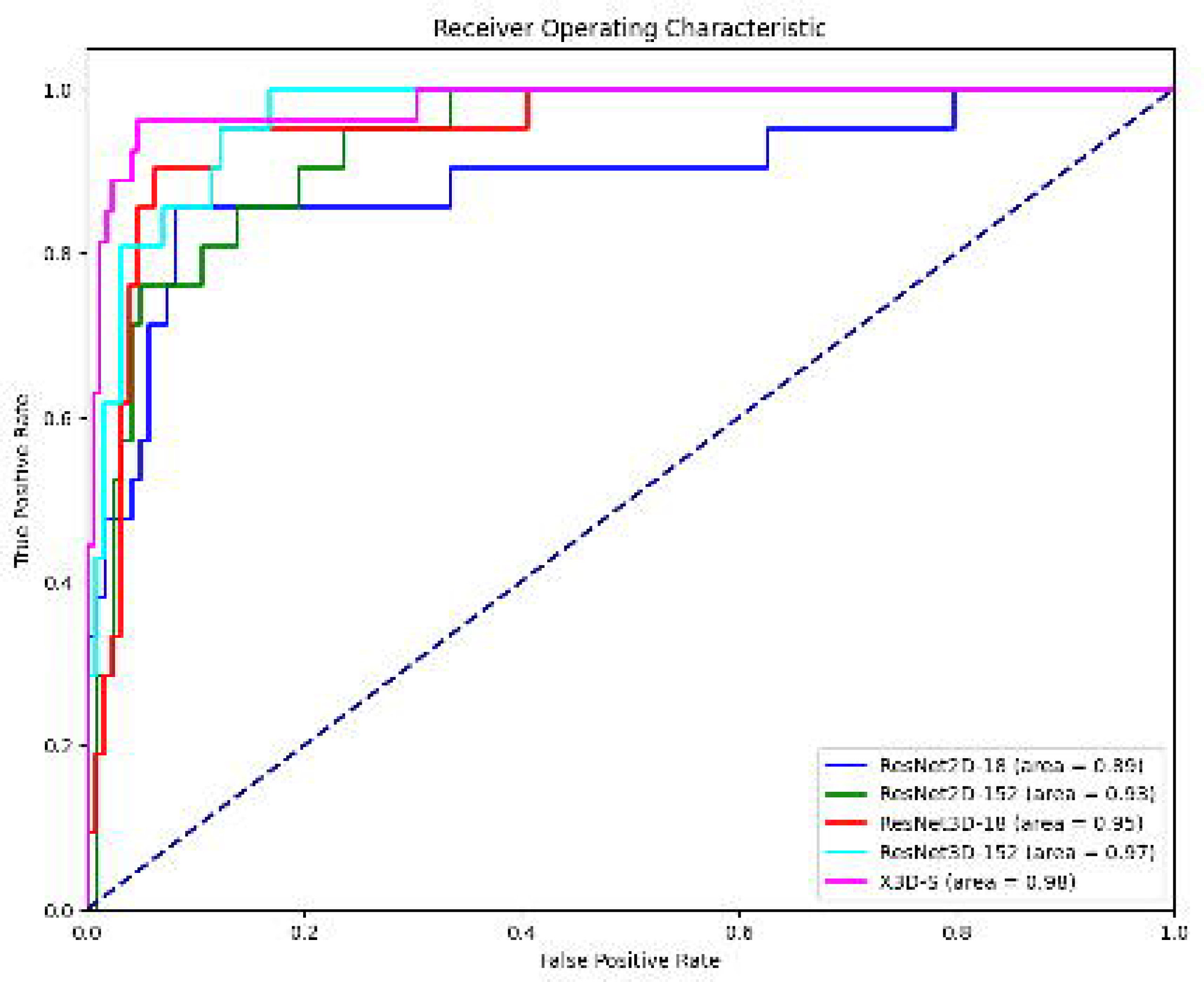
Comparative Analysis of AUC Scores Across 2D and 3D CNN Models for Image Quality Classification.

**Figure 2.**
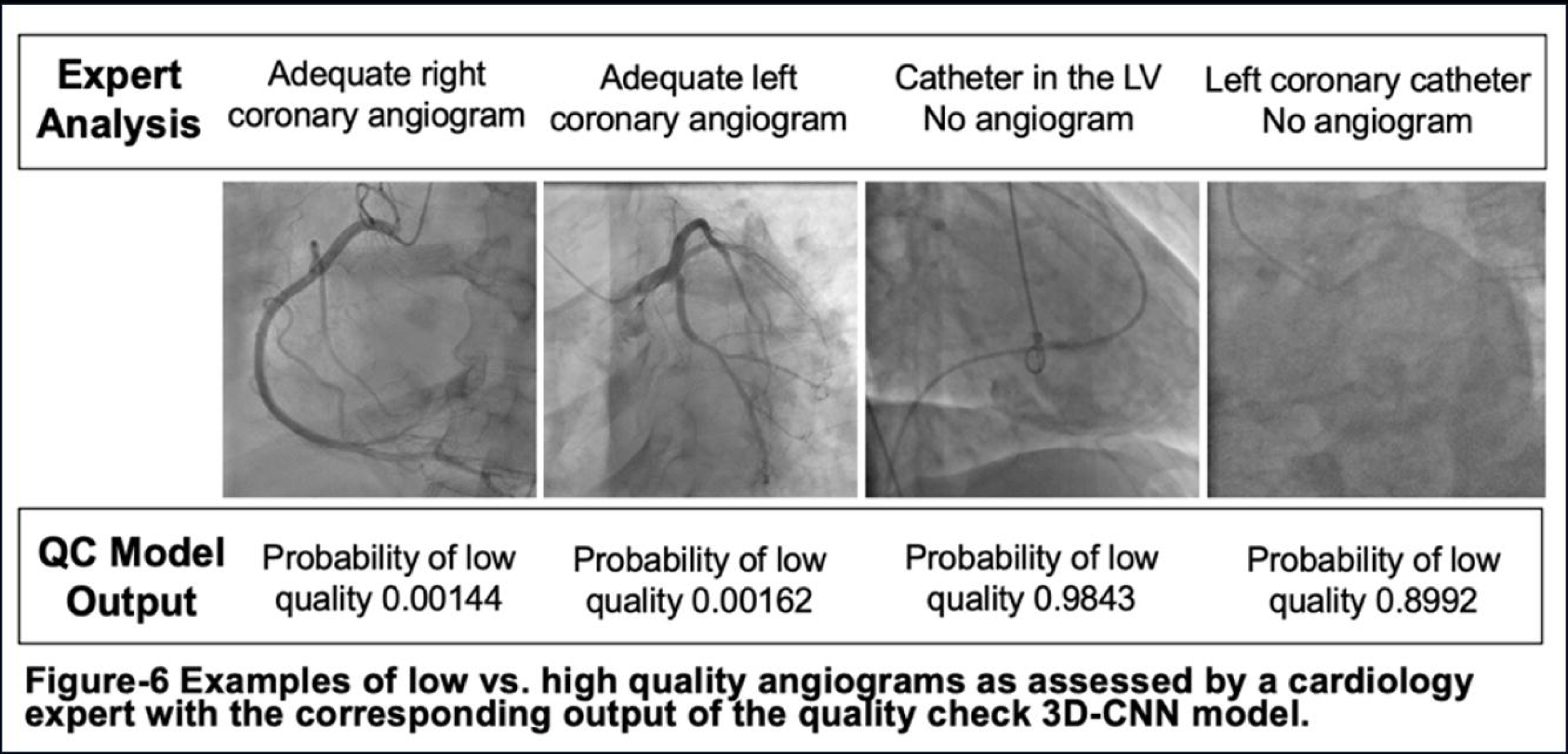
Examples of Unsatisfactory Vs. Satisfactory Angiograms as Assessed by Cardiology Experts With A Corresponding Output of The Quality Check 3D-CNN Model.

**Table 2.** Comparative Performance Metrics of CNN Models.

|  | AUC | Accuracy | Precision | Sensitivity | Specificity | F1-score |
| --- | --- | --- | --- | --- | --- | --- |
| ResNet 18 2D | 0.89 | 0.85 | 0.50 | 0.86 | 0.85 | 0.63 |
| ResNet 152 2D | 0.93 | 0.88 | 0.57 | 0.81 | 0.89 | 0.67 |
| ResNet18 3D | 0.95 | 0.87 | 0.51 | **0.95** | 0.85 | 0.67 |
| ResNet 152 3D | 0.97 | 0.88 | 0.54 | 0.90 | 0.88 | 0.68 |
| X3D-S | **0.98** | **0.96** | **0.86** | 0.89 | **0.98** | **0.87** |

Abbreviations: CNN; convoluted neural network, AUC; area under the curve.

### Computational Analysis of CNN Models

The 2D CNN models, ResNet-18 and ResNet-152 exhibited parameter sizes of 11.18 M and 58.15 M, with computational complexities of 3.41GMAC and 21.46 GMAC, respectively. Both models achieved similar frame rates of approximately 164.5 fps and 207.1 fps, with a processing time of 24.31 and 19.32 millisecond (ms) per study indicating their efficiency in handling 2D image data. Transitioning to 3D architectures, ResNet-18 and ResNet-152 resulted in a significant increase in parameter sizes to 33.2 M and 117.41 M while the computational complexities rose to 59.79 GMAC and 136.11 GMAC, respectively. These models processed the data at the rates of 268.8 fps and 239.2 fps, where each study taking 67.99 and 66.90 ms, reflecting the increased computational demand for 3D image analysis. Notably, the X3D-S model, maintained a balance between computational efficiency and performance with a minimal parameter size of 2.98 M and a computational complexity of 2.56 GMAC. It achieved a processing speed of 267.4 fps and took 59.83 ms per study, highlighting its capability to deliver high performance with less resource utilization as compared to other 3D models. This efficient performance underscores the potential of X3D-S in applications requiring high-speed and accurate 3D image analysis with constrained computational resources.

### Validation of 3D CNNs on an Independent Dataset

Further evaluation of the 3D CNN models on an independent dataset for cardiac function analysis included 20,388 video clips from 2,725 patients. The performance metrics outlined in **Table 4** reveal that the models performed similarly well. The X3D-S model slightly edged out with an AUC of 0.90, accuracy of 0.88, sensitivity of 0.78 and specificity of 0.89, respectively, despite its precision being lower at 0.49. The ResNet-18 3D and ResNet-152 3D models achieved an AUC of 0.90 and 0.88, with comparable sensitivity and specificity values, illustrating their robustness in assessing the quality of angiographic video clips across a diverse dataset. The confusion matrix in **Figure 3** illustrates that the X3D model correctly predicted 1,920 unsatisfactory and 15,971 satisfactory angiograms while misclassifying 1,966 satisfactory and 531 unsatisfactory cases, indicating a reliable quality assessment with a limited misclassification rate.

**Figure 3.**
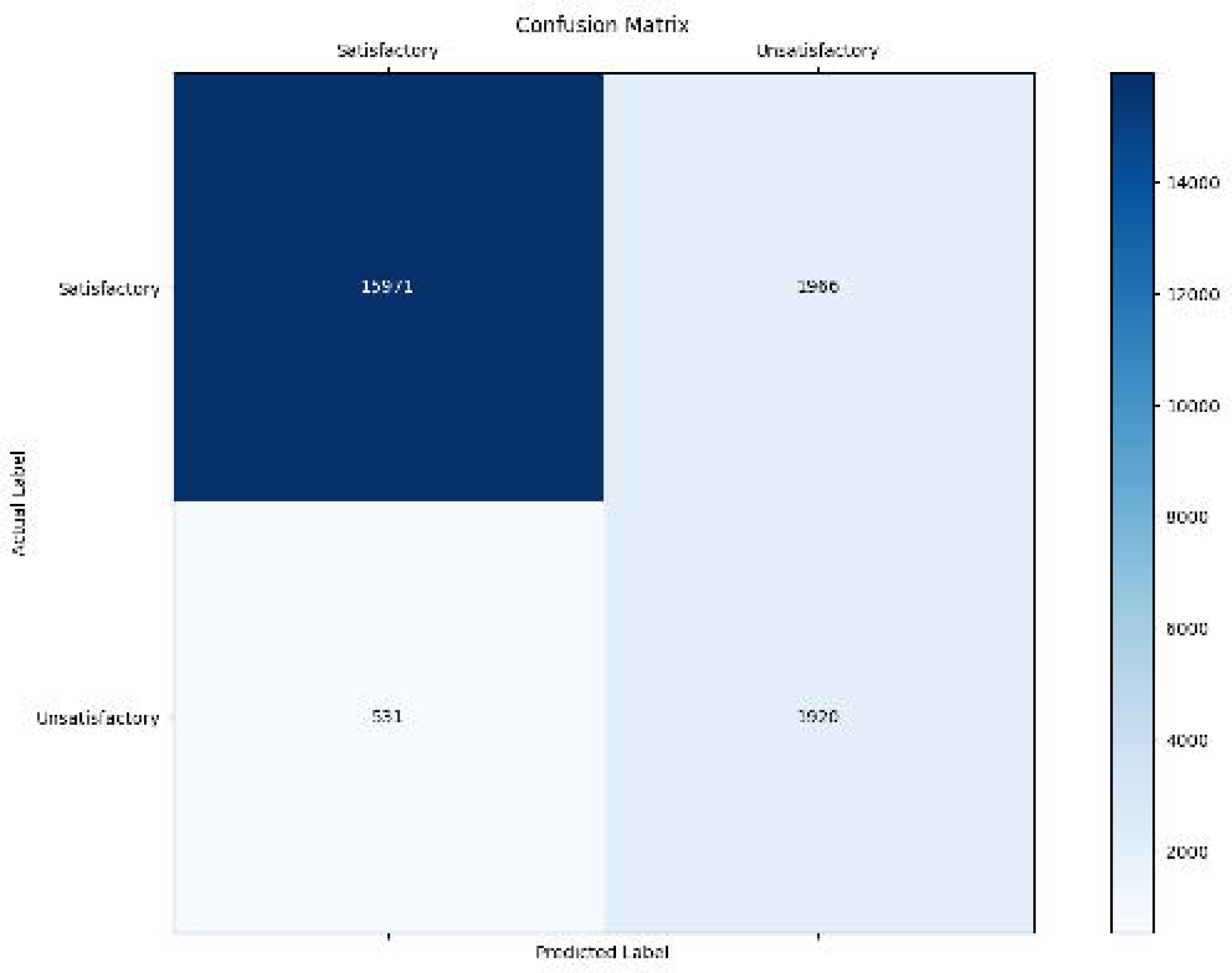
Confusion Matrix of The X3D Model Applying The Independent Dataset.

**Table 3.** Model Complexity and Computational Efficiency.

|  | **Parameters (M)** | **MAC (G)** | **FPS** | **Processing Time**<br>**Per Clip (ms)** |
| --- | --- | --- | --- | --- |
| ResNet 18 2D | 11.18 | 3.41 | 164.5 | 24.31 |
| ResNet 152 2D | 58.15 | 21.46 | 207.1 | 19.32 |
| ResNet18 3D | 33.2 | 59.79 | 268.8 | 67.99 |
| ResNet 152 3D | 117.41 | 136.11 | 239.2 | 66.90 |
| X3D-S | 2.98 | 2.56 | 267.4 | 59.83 |

Abbreviations: M, million; FLOPs, floating-point operations per second; MAC, multiply-accumulate operations; G, giga; FPS, frames per second; ms, millisecond.

**Table 4.** Predictive Outcomes on Independent Dataset.

|  | **AUC** | **Accuracy** | **Precision** | **Sensitivity** | **Specificity** | **F1-score** |
| --- | --- | --- | --- | --- | --- | --- |
| X3D-S | **0.90** | **0.88** | **0.49** | **0.78** | **0.89** | **0.61** |
| Res3D-18 | **0.90** | 0.87 | 0.48 | **0.78** | 0.88 | 0.59 |
| Res3D-152 | 0.88 | 0.86 | 0.44 | 0.74 | 0.87 | 0.56 |

Abbreviations: AUC, area under the curve.

## Discussion

This research introduces an innovative and efficient deep learning approach for automating the selection of appropriate X-ray angiogram videos for ML-driven research in the Cath lab. It features a video selection method that benefits both the training of models and pre-inference screening. When evaluating various CNN models, 3D CNNs—particularly ResNet-152 and X3D-S—emerged as the top performers. Notably, the X3D-S model appears to strike an optimal balance between computational efficiency and complexity, rendering it highly suitable for clinical applications. This method was validated using an independent dataset comprising 20,388 videos from 3,208 angiographic studies, demonstrating the models’ reliability and practical applicability (**Figure. 4**). This advancement offers a significant contribution to cardiac imaging by enabling the automated selection of high-quality coronary angiogram videos, thereby enhancing the accuracy of AI models in these fields.

**Figure 4.**
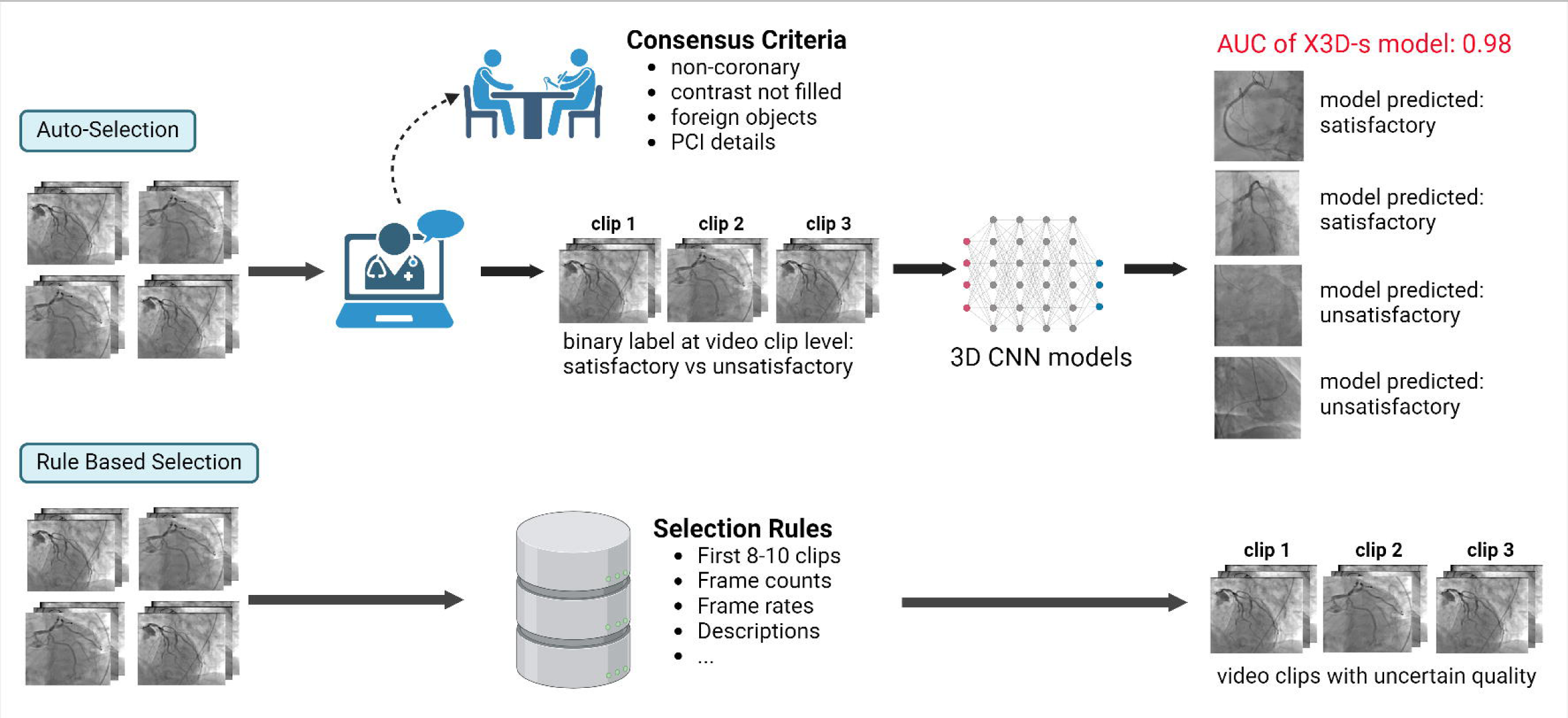
Central Illustration: Workflow of Model Development. PCI; percutaneous coronary intervention, CNN; convoluted neural network, AUC; the area under the receiver operating characteristic curve

There is a limited of literature regarding the auto-selection of suitable video X-ray images for training ML models. This could be attributed to the fact that past studies on angiograms mainly focused on delineating the coronary arteries, while training data has been commonly aligned with frame-by-frame labels. Nonetheless, when the model’s output pertains to cardiac function or predicting clinical events, it is often redundant to meticulously analyze and label each video clip of training data. Rather, each angiographic study is assigned a study-wise label for model training. This necessitates an automated method to help with excluding the inappropriate cine clips from each angiographic exam. A common approach involves setting rules based on information in the DICOM headers, such as selecting the first few clips, ensuring the selection of a minimum number of frames, and specifying a particular frame rate. Other existing automated methods^2,7^, such as keyframe selection and object detection suffer from major limitations that hinder their performance in selecting appropriate videos for input. Consequently, this study introduces an alternative and useful approach to automatically selecting qualified angiographic images for model inputs. This method may simplify the preparation of angiographic data for machine learning applications, thereby facilitating more accurate and efficient model development.

In our study, evaluations were conducted on an independent dataset containing 20,388 video clips from 2,725 patients, examining the ResNet-18 3D, ResNet-152 3D, and X3D-S models. The X3D-S model demonstrated the best performance in terms of AUC and accuracy. Taking the X3D model as an example, its automated screening recommended excluding 19.1% of videos, as opposed to a 12.0% exclusion rate seen with manual review. Of the videos classified as satisfactory by the model, 96.8% conformed to quality standards, indicating a high negative predicted value of the model. However, the application of the X3D-S model to the independent dataset led to a decrease in AUC from 0.95 to 0.90 and a reduction in sensitivity from 0.89 to 0.78. Similar declines in performance were observed in the ResNet-18 3D and ResNet-152 3D models, highlighting the challenge of maintaining consistent performance across different datasets.

The variability in model performance across datasets is significantly influenced by the diverse and complex nature of these datasets, complicating the establishment of a universal standard for classifying images as unsatisfactory. This complexity is further heightened in the case of coronary angiography images from patients with coronary artery bypass grafting, complicating the image selection process. While graft angiograms from patients with prior surgeries were excluded, images where native vessels are clearly visible remained in consideration to preserve essential data. However, the post-surgery appearance of native vessels can vary greatly, leading to potential exclusion without consistent criteria. Moreover, the task of accurately delineating coronary arteries is complicated by the presence of foreign objects in the image, which can obscure crucial details. This adds another layer of complexity to maintaining uniform evaluation standards across different datasets.

### Limitations

This study’s limitations stem from using datasets exclusively from the Mayo Clinic limiting the model’s generalizability without external validation. Despite this, the model performed well across Mayo Clinic’s diverse locations, suggesting institution-specific generalizability might not be critical for image selection in model training. In future studies, fostering inter-institutional collaborations to gather cardiac catheterization X-ray images from varied regions and populations could significantly broaden the model’s applicability. Moreover, leveraging transfer learning^18^ strategy or using attention-based mechanisms^19^ may improve the model’s adaptability and precision effectively.

## Conclusion

This research presents a streamlined and efficient methodology for creating an automated 3D CNN-based quality evaluation tool for video coronary angiograms, exhibiting significant accuracy and effectiveness. The principal advantage of this model is its facilitation of accelerated machine learning model development within Cath lab research domains, particularly for studies necessitating extensive video angiogram analysis. Furthermore, it possesses the potential to automate input selection for models in real-world applications, thereby augmenting predictive precision.

## Data Availability

All data produced in the present study are available upon reasonable request to the authors.

## Notes

### Competing Interest Statement

The authors have declared no competing interest.

### Funding Statement

This study did not receive any funding

### Author Declarations

Ethics committee/IRB of Mayo Clinic gave ethical approval for this work.

